# Examining and classifying reasons for missing viral loads among adults living with HIV: An extended outcome investigation and ascertainment approach in Western Kenya

**DOI:** 10.1101/2024.11.22.24317791

**Authors:** Harriet Fridah Adhiambo, Jayne Lewis-Kulzer, Edwin Nyagesoa, Sarah Gimbel, Eliud Akama, Dorothy Mangale, Lina Montonya, Ingrid Eshun-Wilson, Sarah Iguna, Everlyne Nyandieka, Elizabeth Bukusi, Lisa Abuogi, Thomas Odeny, Maya L. Petersen, Elvin H. Geng

## Abstract

****Introduction**:** Gaps in HIV RNA monitoring persist globally impeding the ability to determine clinical progress and outcomes. This study systematically evaluated provider (e.g., guideline non-adherence), system (e.g., laboratory error) and patient-based (e.g., refusal) drivers of missed viral load (VL) monitoring measurements among adults living with HIV in Kenya.

****Methods**:** Adults aged 18-65 years were followed across five health facilities in Kenya as part of a clinical trial (**NCT#02338739**) where HIV RNA monitoring was done routinely. Instances of missed VL despite being indicated per Kenyan guidelines were identified. An algorithm for assessing root causes of missing HIV RNA was developed and generalized linear models estimated the risk ratios (RR) for patient-level characteristics associated with missed viral load.

****Results**:** Among 1,754 patients (66% female), the prevalence of missed viral load in year one and two was 24.4% and 29.4%, respectively. Drivers for missed viral load measurements included loss to follow up (51.5% in year one and 57.8% in year two), clinician non-adherence with guidelines (36.7% in year one and 32.2% in year two), unknown (10.3% in year one and 8.6% in year two), and requested but not collected (1.5% in year one and 1.3% in year two). Patients aged < 24 years (RR 2.27, 95% CI: 1.66-3.12), those with higher socioeconomic status (RR 1.47, 95% CI: 1.03- 1.91), receiving HIV treatment at a rural clinic (RR 1.22, 95% CI: 1.02-1.46) and with advanced HIV disease (RR 2.39, 95% CI: 1.52-3.73) were more likely to miss VL monitoring.

****Conclusions**:** Missed routine viral load monitoring remains high, primarily due to loss to follow-up, and may substantially alter suppression estimates. Sustainable approaches to keep people living with HIV engaged in care, alongside strengthening providers’ clinical practices and alignment with national guidelines, are necessary for optimizing viral monitoring and accurately assessing viral suppression within public health systems.

## Introduction

The HIV response has greatly reduced mortality globally and in sub-Saharan Africa, including Kenya [1]. The introduction of routine viral load monitoring for people on treatment for HIV in Kenya was anticipated to be a gamechanger in 2013 [2]. Universal access to viral load allows clinicians to monitor patient health more effectively - detecting treatment failure earlier than laboratory measures such as CD4 cell counts or clinical symptoms such as opportunistic infections, resulting in improved management and decreased morbidity and mortality [3].

Previously viral load measurements were reserved for complex patient cases or suspected viral cases among extremely ill patients, with routine care measures relying on clinical or immunological indicators [3], [4].

Despite the importance of routine viral load monitoring, gaps in HIV RNA monitoring persist, impeding clinician ability to determine and shape patients’ clinical progress and outcomes.

Existing data suggest that missed opportunities for viral load monitoring stem from a variety of complex and multilevel health system barriers including reagent stock outs, staff shortages, poor equipment maintenance and delayed clinical review and action on viral load results all of which impede progress toward epidemic control [5], [6]. An inefficient viral load monitoring system not only compromises a clinician’s ability to accurately detect treatment failure, but also delays provision of timely treatment interventions to patients experiencing virologic failure, leading to increased morbidity (including drug resistance), risk of HIV transmission as well as risk of mortality [7], [8]. Additionally, there is a risk of missing broader trends and accurately measuring viral suppression within the population.

Research has focused on associated factors of virologic failure, viral load uptake and monitoring, and cost effectiveness of routine viral load monitoring [9], [10], [11], [12], [13]. The WHO recommends routine viral load testing at 6 and 12 months post ART initiation and every 12 months thereafter [14]. In Kenya, although routine viral load monitoring has improved since its introduction, patient follow up and data management systems at the facility level are known barriers to the viral load monitoring process [15]. However, there is limited data examining the degree of drivers of missing viral load among people with HIV [8]. To better discern issues contributing to these gaps, we developed an algorithm to assess the root causes of missing HIV RNA among adult patients with HIV on antiretroviral therapy (ART) and examined patient characteristics and processes associated with timely viral load monitoring.

## Methods

### Study Design

This cohort study was nested within a larger sequential multiple assignment trial (SMART), AdaPT-R (NCT#02338739) study in Kenya. In the larger trial, HIV RNA monitoring during the study was conducted entirely through routine service delivery processes at public health facilities (i.e., the research staff did not ensure monitoring) and were thus representative of wider health systems practice.

### Study setting

The AdaPT-R study was conducted in the Nyanza region of western Kenya from September 2014 to December 2019 at five health facilities, including four government hospitals and one faith-based health center [16]. Three sites were in urban Kisumu County: Lumumba Sub County Hospital, Ahero Sub County Hospital, and Pandipieri Health Center. The other two sites were in more rural areas including Migori County: Rongo Sub County Hospital and Migori County Referral Hospital. Both counties have a high HIV prevalence, 19.3% in Kisumu and 13.3% in Migori, compared to the national prevalence of 4.9% [17], [18], [19].

### Study population

The study population included people living with HIV (PWH), ≥ 18 years of age who had initiated ART within 90 days of study enrollment, planned to remain in the Nyanza region for the duration of the study, had cell phone access and ability to read or be read short messages (SMS) messages, and were willing to be contacted by a clinic worker upon missing a clinic appointment. Participants were recruited and enrolled from HIV clinics at the five health facilities. In this sub-study, we included all AdaPT-R participants. Participants who missed viral load measurements during their 1^st^ and 2^nd^ year in the study. AdaPT-R participants that had died or withdrew from the study before the extended outcome investigation were excluded.

### Ethical approvals

The institutional review boards of the University of California, San Francisco (UCSF IRB No. 13–12810) and the Kenya Medical Research Institute (KEMRI SSC No 2838) in Kenya approved this study. All participants provided informed consent to participate.

### Measurement of patient characteristics associated with missed viral load

The sociodemographic, socioeconomic, and clinical characteristics of participants were captured at study enrollment from March 2015-October 2018. Data was entered in Open Data Kit (ODK) and the patient clinical characteristics, collected as part of routine patient care, were entered into an electronic medical record system maintained by the Family AIDs Care and Education

Program (FACES). The data from both sources were then synchronized daily to the study database. For this sub-study, sociodemographic variables of interest were abstracted from the study database at study enrollment and included age, gender, educational status, marital status and socioeconomic variables including household size, and occupation, as well as clinical characteristics including WHO stage, baseline CD4, study site, and viral load history.

### Procedures and Measurement

This study and the participating health facilities followed National ART Guidelines for viral load monitoring which recommend that viral load testing be conducted 6- and 12-months post ART initiation and annually thereafter for adults with viral suppression (defined as <1000 copies per mL during the study period).[20] Patients without viral suppression undergo adherence counseling and repeat viral load is recommended after three months of good adherence. As part of routine care, patient viral load samples are taken at the facility and then sent to centralized laboratories including the Kenya Medical Research Institute -Center for Disease Control (KEMRI-CDC) laboratories at Kisian in Kisumu and the Academic Model Providing Access to Healthcare laboratories (AMPATH) in Eldoret for processing. As part of national standards, all results are uploaded and stored on the National STI and AIDs Control Program (NASCOP) website by the laboratories and results are returned to the health facility. Results are then updated in patient medical records, ideally both in paper files and in the electronic medical system (EMR). Patients are then informed of results during their next clinic visit if they are suppressed and notified to return to the clinic for their results if they are not suppressed. To examine viral load missingness and drivers of missingness in this sub-study, a two-step process was employed.

#### **<u>Step 1</u>**: Outcome ascertainment (confirming missingness)

The study identified patients who did not have a HIV RNA measurement documented in the study database between 9-15 months post ART initiation and 21-27 months post ART initiation (i.e., windows in which guideline recommended 12- and 24-month HIV RNA measurements are to be obtained). Research assistants then explored clinic records by checking the EMR, the patient paper medical records, and the NASCOP website for any viral load within the established window period. If a viral load measurement was found during the window period, the result and date were documented on the report and entered in the study database; the participant was then removed from the missing viral load report. Those who remained on the missing viral report moved to Step 2.

#### **<u>Step 2:</u>** Ascertainment of reason for missing viral load

This involved investigating the missing viral load measurement further through an “extended investigation” and, if the HIV RNA was confirmed missing, classifying the reason for missingness. This occurred during the same month as step 1 then continued monthly until all reviews were completed for a single participant. Reasons for missingness were classified based on reviews of patient records and consultations with facility clinicians through a process which encompassed research assistants conducting a series of review in the following order: 1. In-depth review of the entire patient file; 2. Repeat review of the EMR; 3. Examination of laboratory records and logs. 4. Repeat review of the NASCOP website; 5. Review study records (e.g., tracking logs); 6. Consultation with research assistants for participants in their case load; and 7. Review study lost to follow up records. Viral loads found through this extended investigation were documented in the study database and the participant was removed from missingness report. Classification of missingness was further informed by patient status during the window period of interest and included death, official or self-transferred to another facility, and non-transfer (Appendix 1). If a non-transfer, the patient file was reviewed to see if the patient had a clinic visit during the viral load window period of interest and if they did, clinical factors were examined to determine if it was a: 1. Clinician decision and 2. Viral load ordered, but sample not collected. The patient status outcomes and clinical factor outcomes were entered in the study database.

### Analysis

Missed viral load was defined as no viral load results within 9-15 months (First year of study) and 21-27months of ART initiation (Second year of study). Descriptive statistics were used to determine the prevalence of missed viral load and reasons for missing viral load measurements. In our analysis we conducted complete case analyses by excluding missing values.

Socioeconomic status (SES) indices were generated using multiple correspondence analysis (MCA) using the following variables: occupation of household head, primary source of drinking water, type of cooking fuel, ownership of household assets and ownership of livestock. The households were categorized into five socioeconomic quintiles classified as poorest, poorer, middle, richer and richest [21], [22]. A generalized linear model, using a Poisson distribution with a log-link function, was used to estimate adjusted risk ratios (RR) to determine the patient-level characteristics associated with missed viral load in year one and year two [23]. We identified covariates from the literature review and used them in the bivariate and multivariate analysis [24], [25], [26].

## Results

A total of 1815 participants were enrolled to the Adapt-R study. **Table 1** below describes participant characteristics at one and two years of follow-up. Sixty-six percent of study participants were females and over half (54.8%) were from urban clinics. No differences in background characteristics in the first and second year.

**Table 1:**
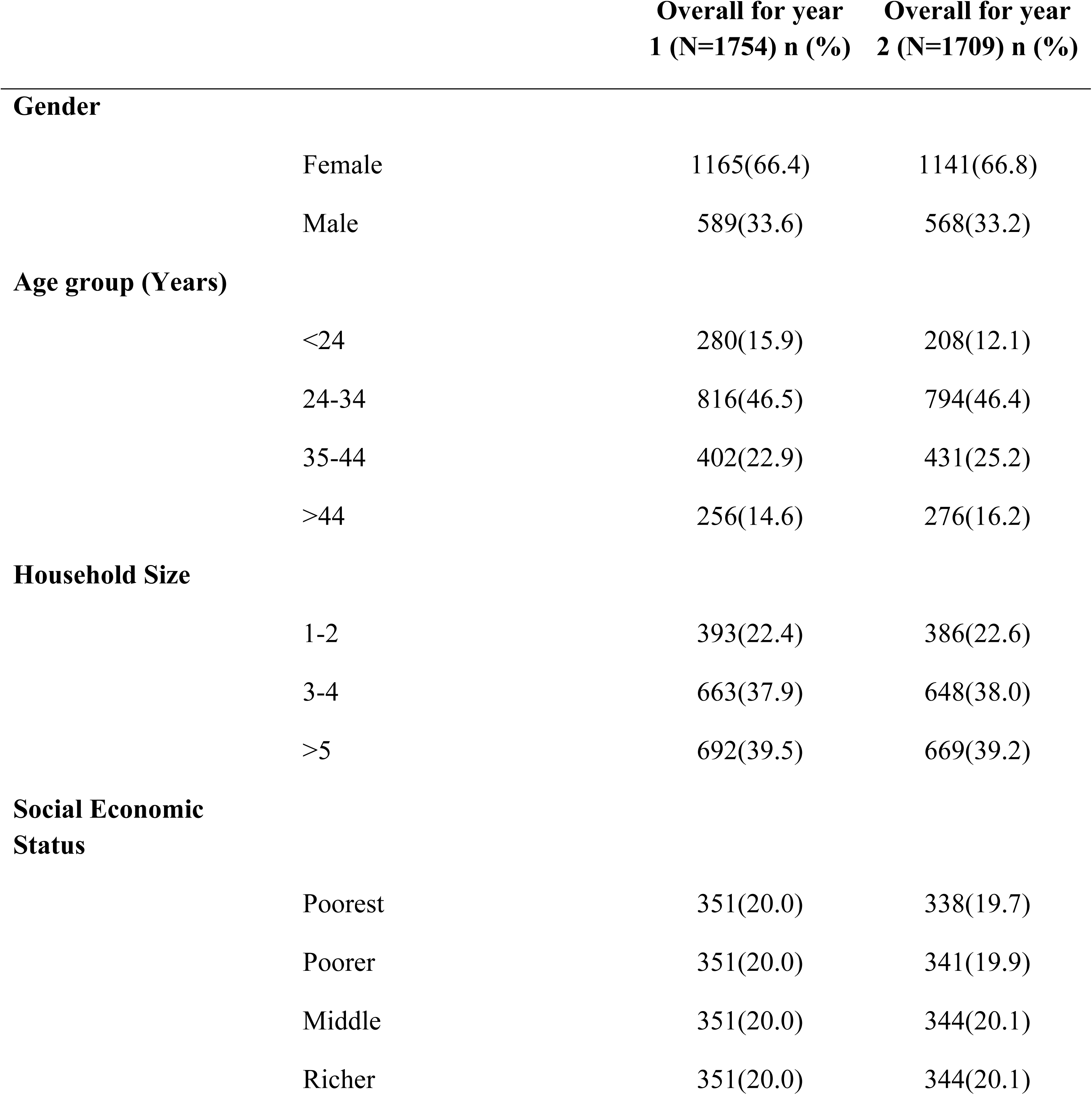
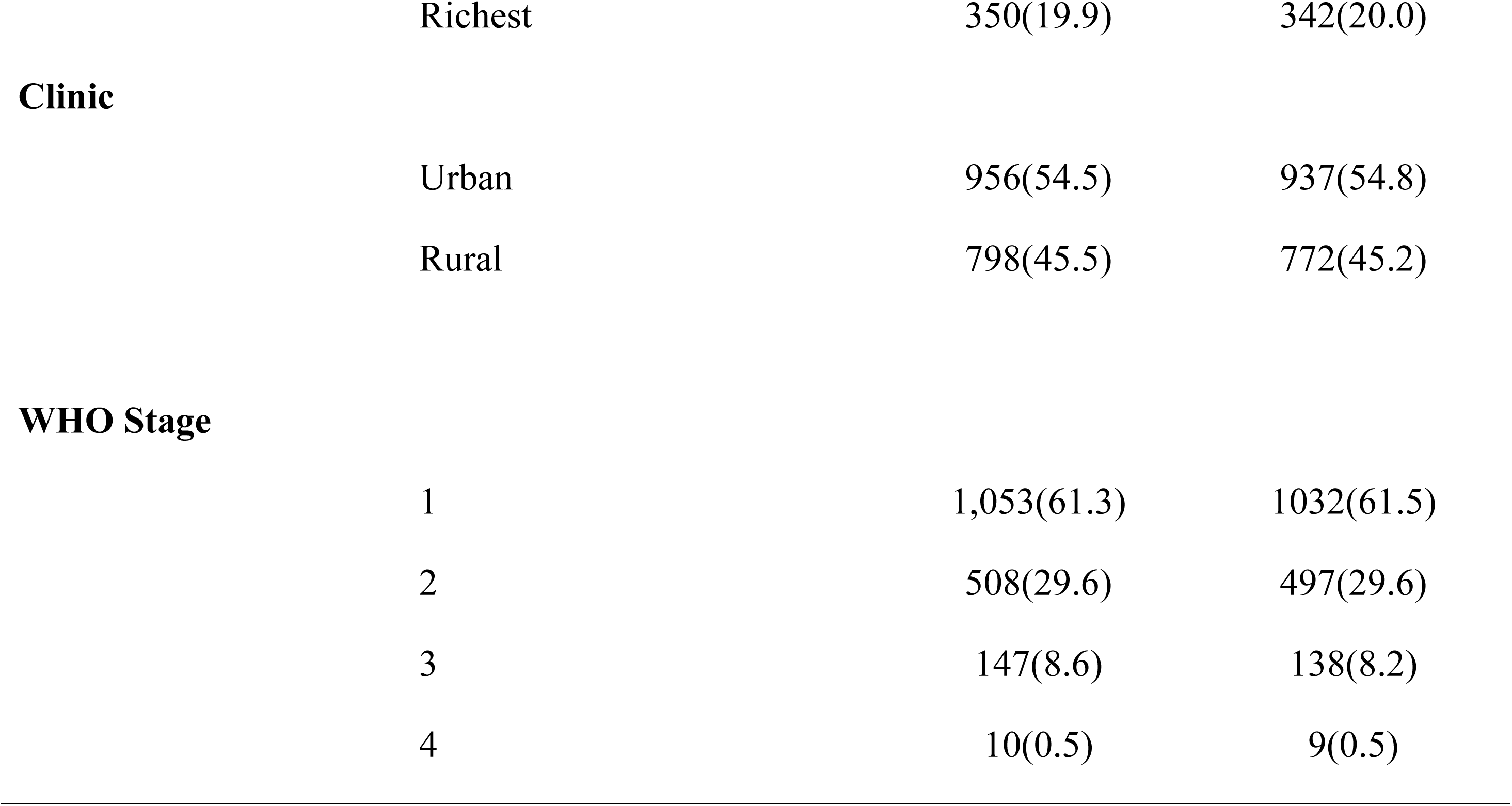
Background Characteristics

|  |  | **Overall for year**<br>**1 (N=1754) n (%)** | **Overall for year**<br>**2 (N=1709) n (%)** |
| --- | --- | --- | --- |
| **Gender** |  |  |  |
|  | Female | 1165(66.4) | 1141(66.8) |
|  | Male | 589(33.6) | 568(33.2) |
| **Age group (Years)** |  |  |  |
|  | <24 | 280(15.9) | 208(12.1) |
|  | 24-34 | 816(46.5) | 794(46.4) |
|  | 35-44 | 402(22.9) | 431(25.2) |
|  | >44 | 256(14.6) | 276(16.2) |
| **Household Size** |  |  |  |
|  | 1-2 | 393(22.4) | 386(22.6) |
|  | 3-4 | 663(37.9) | 648(38.0) |
|  | >5 | 692(39.5) | 669(39.2) |
| **Social Economic**<br>**Status** |  |  |  |
|  | Poorest | 351(20.0) | 338(19.7) |
|  | Poorer | 351(20.0) | 341(19.9) |
|  | Middle | 351(20.0) | 344(20.1) |
|  | Richer | 351(20.0) | 344(20.1) |
|  | Richest | 350(19.9) | 342(20.0) |
| **Clinic** | Urban | 956(54.5) | 937(54.8) |
|  | Rural | 798(45.5) | 772(45.2) |
| **WHO Stage** | 1 | 1,053(61.3) | 1032(61.5) |
|  | 2 | 508(29.6) | 497(29.6) |
|  | 3 | 147(8.6) | 138(8.2) |
|  | 4 | 10(0.5) | 9(0.5) |

### Prevalence of missed viral load

Prevalence of missing viral load at year one was (428/1754) 24.4% whereas the prevalence of missing viral load at year two was (502/1709) 29.4%. The cumulative incidence of viral load monitoring was 72.3% and 68.9% in year one and two respectively (Table 2) with an additional 15.6% at year one and 7.1% at year two having delayed viral loads measured.

**Table 2:**
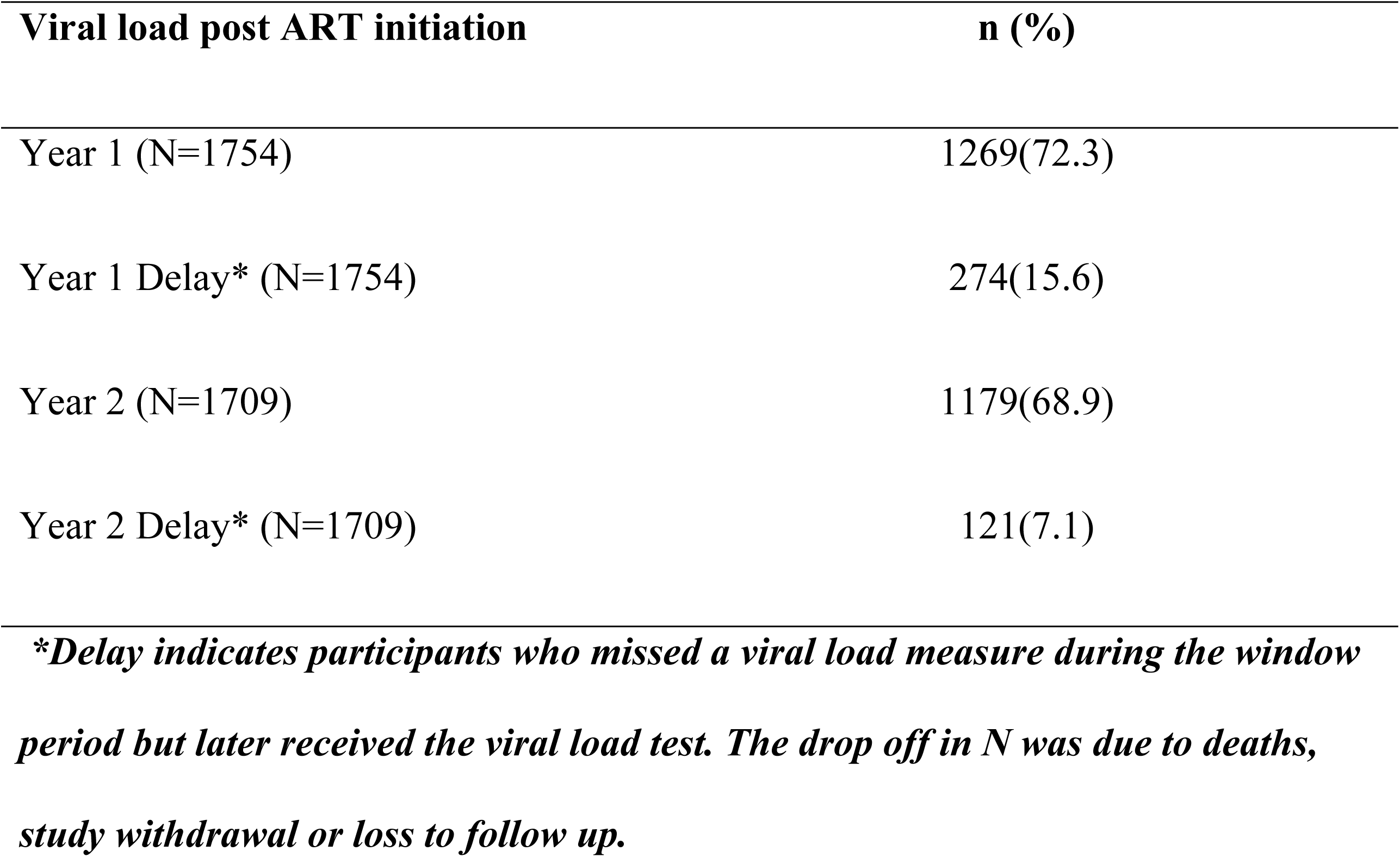
Cumulative incidence of viral load monitoring among ADAPT-R study participants

| **Viral load post ART initiation** | **n (%)** |
| --- | --- |
| Year 1 (N=1754) | 1269(72.3) |
| Year 1 Delay* (N=1754) | 274(15.6) |
| Year 2 (N=1709) | 1179(68.9) |
| Year 2 Delay* (N=1709) | 121(7.1) |

*\*Delay indicates participants who missed a viral load measure during the window period but later received the viral load test. The drop off in N was due to deaths, study withdrawal or loss to follow up.*

At year one, the most common reason for missed viral load was due to missing all appointments, and thus being lost to follow up, during the VL window period (N= 209 (51.5%). However, of the 406 patients who were alive and missed VL monitoring measurements, 149 (36.7%) missed viral load monitoring as a result of mis-interpretation of routine viral load monitoring guidelines with clinicians ordering VL 12 months from the last VL done as opposed to 12 months from ART initiation., An additional 42 (10.3%) patients had unknown reason for missed VL and 6 (1.5%) patients had viral load ordered but there was no evidence the samples were collected.

Similarly, at year two, of the 475 patients who were alive and did not have a VL measured, the most common reason was missed appointments (N=275 (57.8%)), with 153 (32.2%) patients missing their viral load measurements because of misinterpretation of the guideline timing, 41 (8.6%) with no known documented reason, and 6 (1.3%) had their viral loads requested by the clinicians but there was no evidence the samples were ever collected.(**Figure 2)**

**Figure 1.**
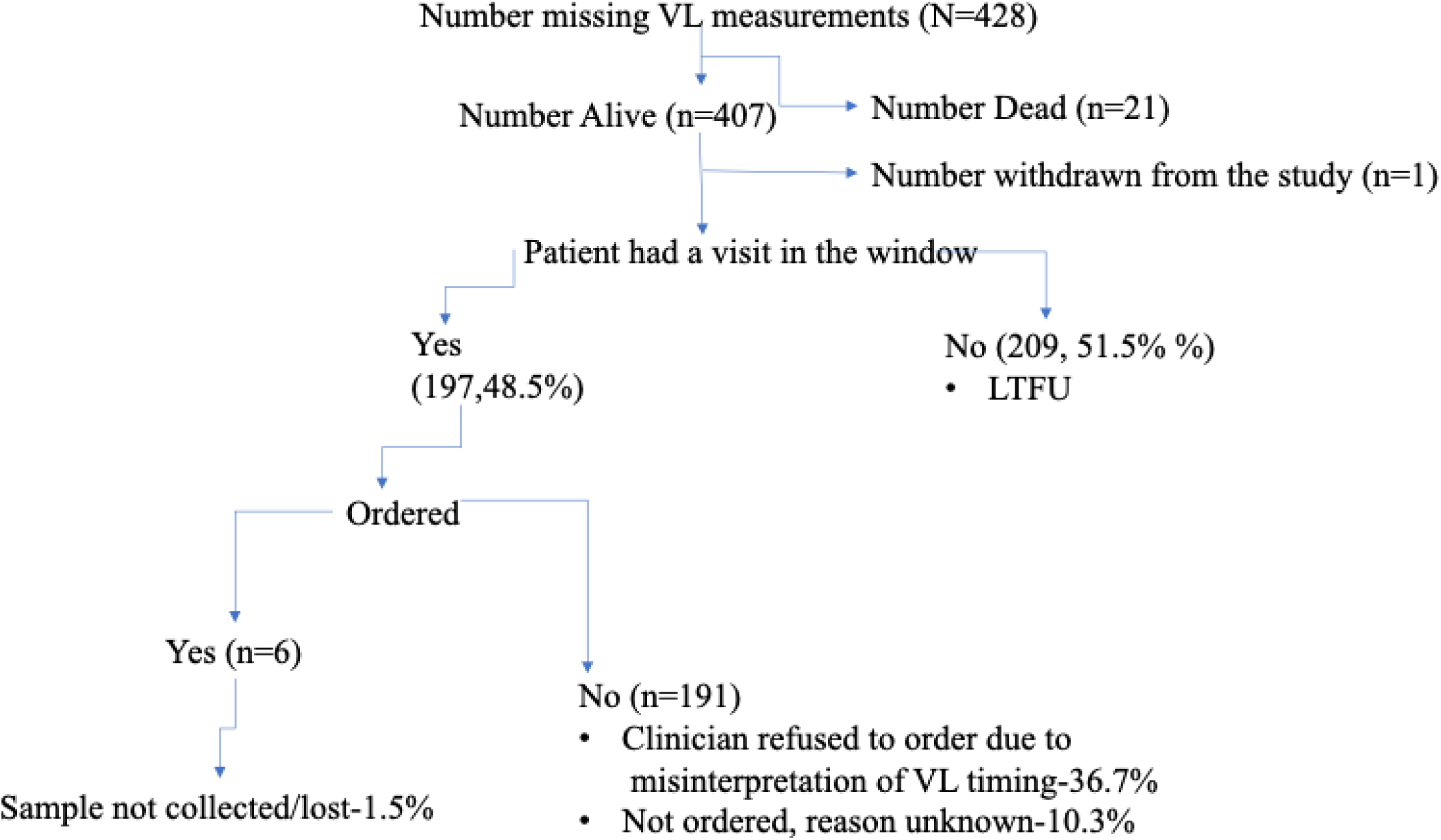
Reasons for Missing Viral Load Measurements at Year 1

**Figure 1a.**
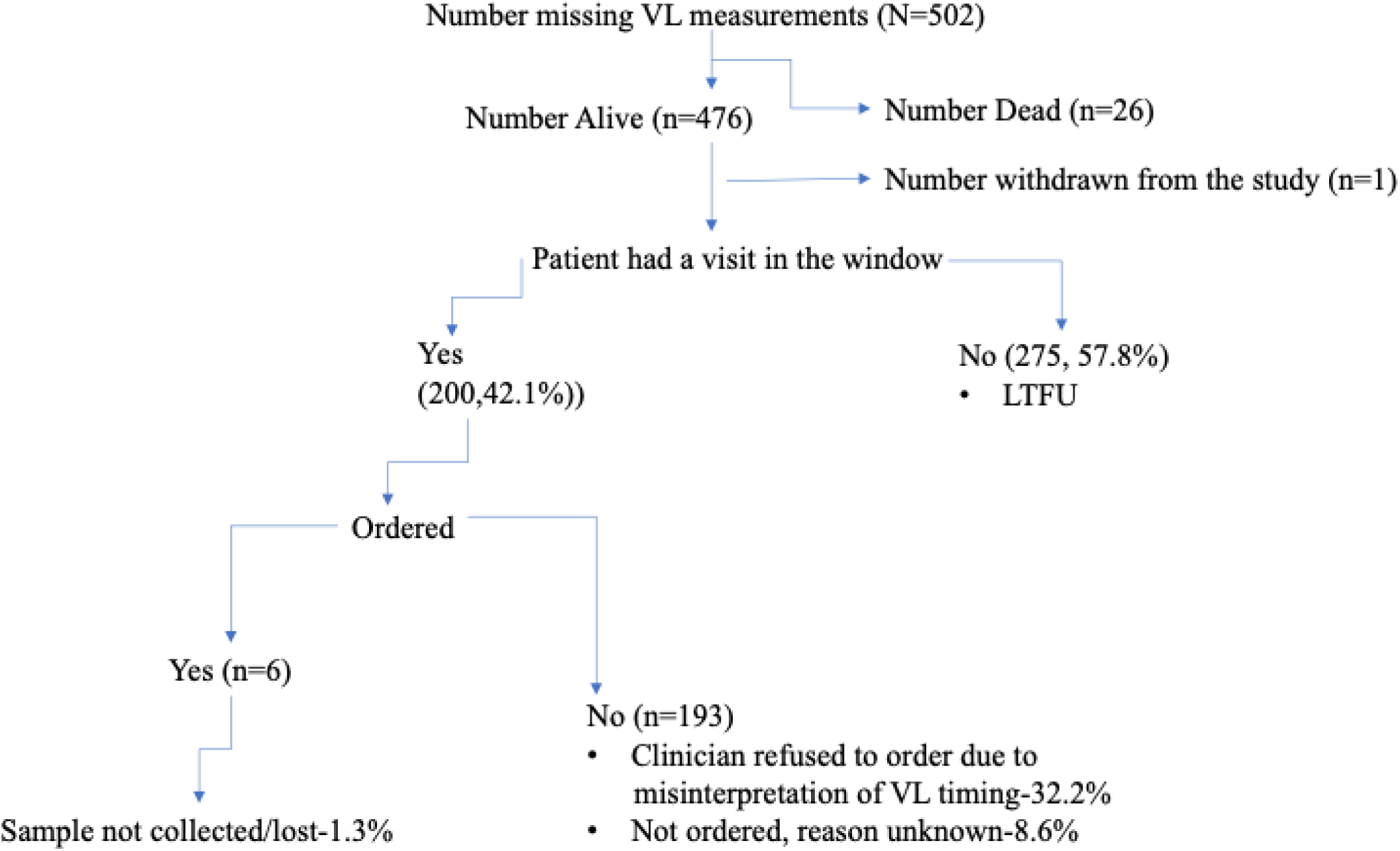
Reasons for Missing Viral Load Measurements at Year 2

Multivariate analysis of patient characteristics associated with missed viral load in year 1 and year 2 reveals that younger participants age < 24 years (Year 1 aRR=2.27;95%CI 1.66-3.12; p<0.001, Year 2 aRR= 1.69; 95%CI 1.28-1.22 p<0.001) had a higher risk of missed viral load as compared to those whose age group is > 44 years. Participants whose Socio-Economic Status (SES) were richer (Year 1 aRR=1.51;95%CI 1.14-1.99; p=0.003 and Year 2 aRR= 1.47 95%CI (1.03-1.91, p=0.004) also had a higher risk of missed viral load as compared to those with poorest SES. Additionally, participants from rural clinics in year 1 (aRR=1.22;95%CI 1.02-1.46; p=0.029) had a higher risk of missed viral load as compared to those whose clinics were urban **(Table 3 and Table 4),** whereas participants in Year 2 categorized as WHO stage 4 had a higher risk of missed viral load compared to participants in WHO stage 1(aRR=2.39; 95%CI 1.52-3.73, P<0.001). Stratified analysis comparing patient characteristics by rural vs urban did not change associations with missed viral load in year 1 and 2 respectively.

**Table 3.**
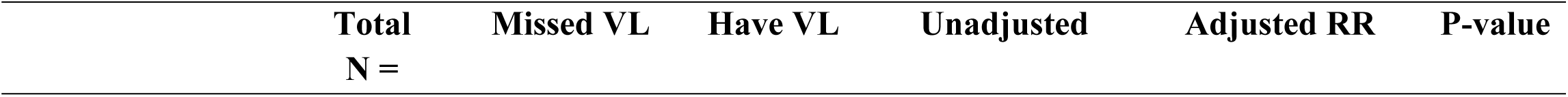
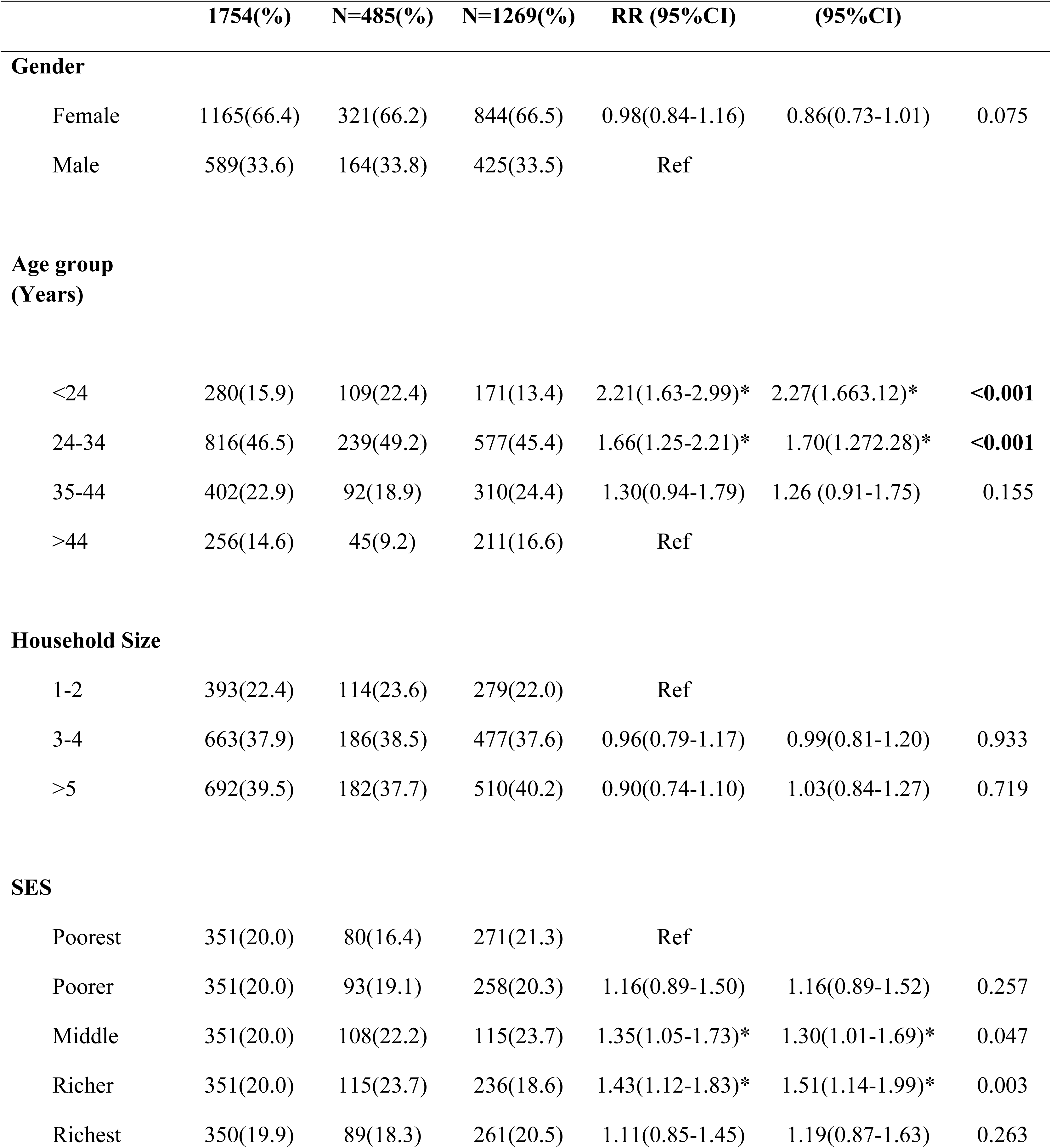
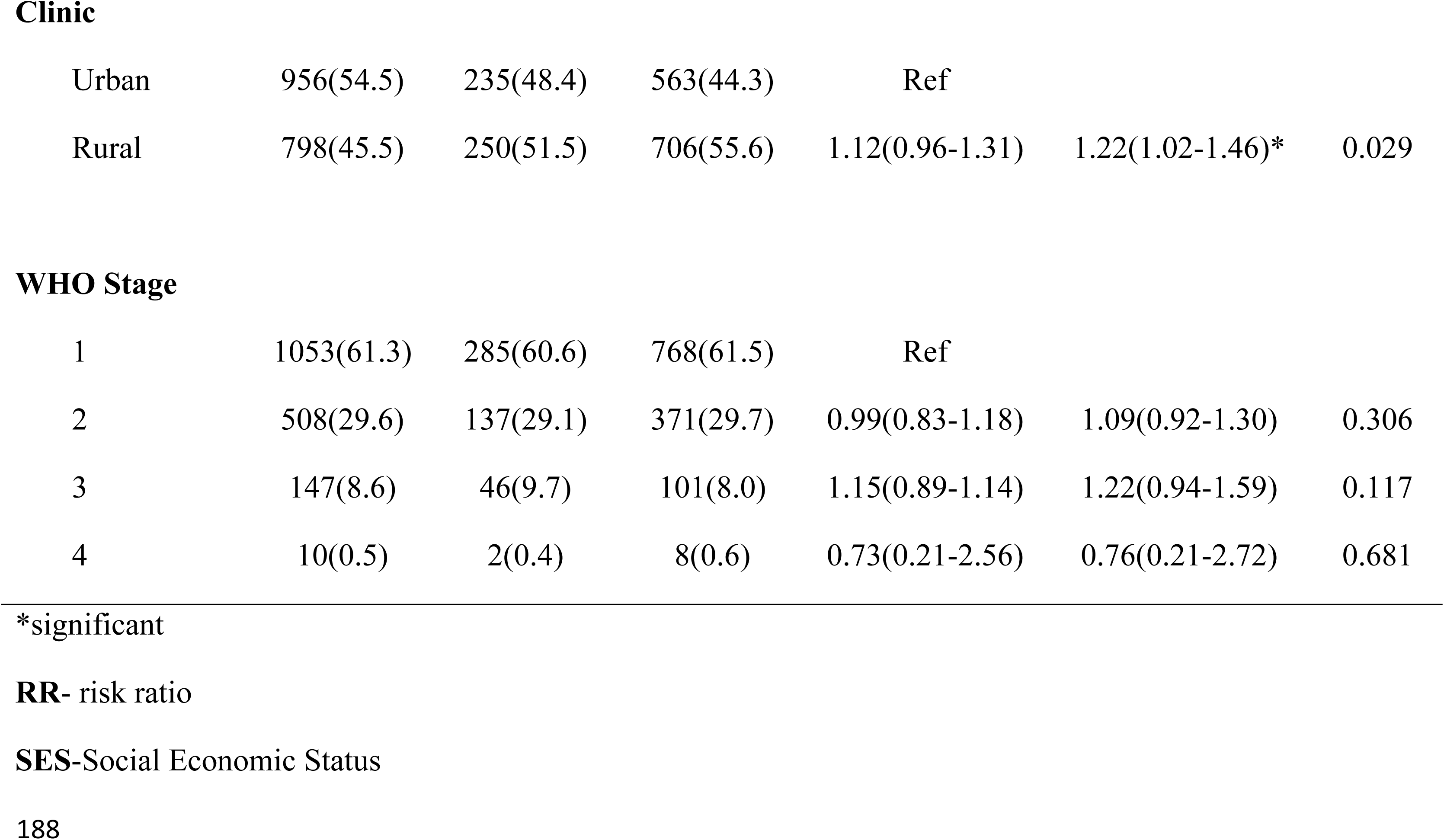
Multivariate analysis of patient characteristics associated with missed viral load in Year1

|  | Total<br>N = | Missed VL | Have VL | Unadjusted | Adjusted RR | P-value |
| --- | --- | --- | --- | --- | --- | --- |

|  | 1754(%) | N=485(%) | N=1269(%) | RR (95%CI) | (95%CI) |  |
| --- | --- | --- | --- | --- | --- | --- |
| **Gender** |  |  |  |  |  |  |
| Female | 1165(66.4) | 321(66.2) | 844(66.5) | 0.98(0.84-1.16) | 0.86(0.73-1.01) | 0.075 |
| Male | 589(33.6) | 164(33.8) | 425(33.5) | Ref |  |  |
| **Age group (Years)** |  |  |  |  |  |  |
| <24 | 280(15.9) | 109(22.4) | 171(13.4) | 2.21(1.63-2.99)* | 2.27(1.663.12)* | <0.001 |
| 24-34 | 816(46.5) | 239(49.2) | 577(45.4) | 1.66(1.25-2.21)* | 1.70(1.272.28)* | <0.001 |
| 35-44 | 402(22.9) | 92(18.9) | 310(24.4) | 1.30(0.94-1.79) | 1.26 (0.91-1.75) | 0.155 |
| >44 | 256(14.6) | 45(9.2) | 211(16.6) | Ref |  |  |
| **Household Size** |  |  |  |  |  |  |
| 1-2 | 393(22.4) | 114(23.6) | 279(22.0) | Ref |  |  |
| 3-4 | 663(37.9) | 186(38.5) | 477(37.6) | 0.96(0.79-1.17) | 0.99(0.81-1.20) | 0.933 |
| >5 | 692(39.5) | 182(37.7) | 510(40.2) | 0.90(0.74-1.10) | 1.03(0.84-1.27) | 0.719 |
| **SES** |  |  |  |  |  |  |
| Poorest | 351(20.0) | 80(16.4) | 271(21.3) | Ref |  |  |
| Poorer | 351(20.0) | 93(19.1) | 258(20.3) | 1.16(0.89-1.50) | 1.16(0.89-1.52) | 0.257 |
| Middle | 351(20.0) | 108(22.2) | 115(23.7) | 1.35(1.05-1.73)* | 1.30(1.01-1.69)* | 0.047 |
| Richer | 351(20.0) | 115(23.7) | 236(18.6) | 1.43(1.12-1.83)* | 1.51(1.14-1.99)* | 0.003 |
| Richest | 350(19.9) | 89(18.3) | 261(20.5) | 1.11(0.85-1.45) | 1.19(0.87-1.63) | 0.263 |
| Clinic |  |  |  |  |  |  |
| Urban | 956(54.5) | 235(48.4) | 563(44.3) | Ref |  |  |
| Rural | 798(45.5) | 250(51.5) | 706(55.6) | 1.12(0.96-1.31) | 1.22(1.02-1.46)* | 0.029 |
| WHO Stage |  |  |  |  |  |  |
| 1 | 1053(61.3) | 285(60.6) | 768(61.5) | Ref |  |  |
| 2 | 508(29.6) | 137(29.1) | 371(29.7) | 0.99(0.83-1.18) | 1.09(0.92-1.30) | 0.306 |
| 3 | 147(8.6) | 46(9.7) | 101(8.0) | 1.15(0.89-1.14) | 1.22(0.94-1.59) | 0.117 |
| 4 | 10(0.5) | 2(0.4) | 8(0.6) | 0.73(0.21-2.56) | 0.76(0.21-2.72) | 0.681 |

\*significant
**RR-** risk ratio
**SES-**Social Economic Status

**Table 4.** Multivariate analysis of patient characteristics associated with missed viral load in year 2

|  | Total<br>N =<br>1709(%) | Missed<br>VL<br>N=520(%) | Have VL<br>N=1189(%) | Unadjusted RR<br>(95%CI) | Adjusted RR<br>(95%CI) | P-value |
| --- | --- | --- | --- | --- | --- | --- |
| **Gender** |  |  |  |  |  |  |
| Female | 1141(66.8) | 343(65.9) | 798(67.1) | 0.98(0.84-1.16) | 0.86(0.73-1.01) | 0.059 |
| Male | 568(33.2) | 177(34.1) | 391(32.8) | Ref |  |  |
| **Age group (Years)** |  |  |  |  |  |  |
| <24 | 208(12.1) | 91(17.5) | 117(9.8) | 1.85(1.42-2.41)* | 1.69(1.28-1.22)* | <0.001* |
| 24-34 | 794(46.4) | 253(48.6) | 541(45.5) | 1.35(1.06-1.71)* | 1.27(0.99-1.63) | 0.059 |
| 35-44 | 431(25.2) | 111(21.3) | 320(26.9) | 1.09(0.83-1.42) | 1.01(0.75-1.33) | 0.997 |
| >44 | 276(16.1) | 65(12.5) | 211(17.7) | Ref |  |  |
| **Household Size** |  |  |  |  |  |  |
| 1-2 | 386(22.6) | 136(26.3) | 250(21.0) | Ref |  |  |
| 3-4 | 648(38.0) | 207(40.1) | 441(37.1) | 0.90(0.76-1.08) | 0.92(0.77-1.10) | 0.381 |
| >5 | 669(39.2) | 173(33.5) | 496(41.7) | 0.73(0.60-0.88) | 0.82(0.68-1.01) | 0.061 |
| **SES** |  |  |  |  |  |  |
| Poorest | 338(19.7) | 84(16.1) | 254(21.3) | Ref |  |  |
| Poorer | 341(19.9) | 95(18.2) | 246(20.6) | 1.12(0.87-1.44) | 1.08(0.80-1.40) | 0.566 |
| Middle | 344(20.1) | 101(19.4) | 243(20.4) | 1.18(0.92-1.51) | 1.11(0.85-1.44) | 0.429 |
| Richer | 344(20.1) | 131(25.1) | 213(17.9) | 1.53(1.21-1.92)* | 1.47(1.03-1.91)* | **0.004\*** |
| Richest | 342(20.0) | 109(20.9) | 233(19.6) | 1.28(1.01-1.63)* | 1.17(0.87-1.57) | 0.287 |
| **Clinic** |  |  |  |  |  |  |
| Urban | 937(54.8) | 288(55.3) | 649(54.5) | Ref |  |  |
| Rural | 772(45.2) | 232(44.6) | 540(45.4) | 0.97(0.84-1.12) | 1.10(0.92-1.57) | 0.264 |
| **WHO Stage** |  |  |  |  |  |  |
| 1 | 1032(61.5) | 325(64.1) | 707(60.4) | Ref |  |  |
| 2 | 497(29.6) | 131(25.8) | 366(31.3) | 0.83(0.70-0.99) | 0.88(0.74-1.05) | 0.185 |
| 3 | 138(8.2) | 45(8.8) | 93(7.9) | 1.03(0.80-1.33) | 1.06(0.82-1.37) | 0.649 |
| 4 | 9(0.5) | 6(1.2) | 3(0.3) | 2.11(1.32-3.38) * | 2.39(1.52-3.73) * | **<0.001\*** |

\*significant
SES-Social Economic Status

Additional findings from the research team’s perspective revealed that some clinicians deliberately avoiding conducting viral load measurements from patients suspected to be viremic. These were patients who reported missing doses of ART or had tendencies of missing their clinic appointments. However, this practice was not documented in the patients file but reported by some clinicians when contacted by the research staff during the extended outcome investigation.

## Discussion

An effective viral load monitoring system is critical to early intervention and prevention of poor health outcomes as a result of viremia for PWH. Poor coverage of clinical monitoring systems impedes the ability of HIV programs and country health officials to determine population outcomes. This study showed that about a third of study participants eligible and due for viral load measurement after initiating treatment were missing their viral load at key monitoring timepoints.

These findings are similar a study conducted in South Africa that showed 32% and 26% of the participants did not get a viral load test in the first and second years after ART start [27], [28]. Although WHO recommendation for viral testing led to rapid scale up of viral load monitoring among PWH on treatment, these findings suggest health system issues resulting in less-than-ideal viral load monitoring. Over the past five years there has been considerable improvement in viral load monitoring; however, gaps in viral load monitoring of patients still persist [30]. HIV treatment success is defined by early diagnosis, prompt ART initiation and viral suppression [31]. With over a third of the patients missing routine VL measurements, VL monitoring is incomplete, leading to undetected cases of failure, resistance, and risk of HIV transmission; subsequently undermining attainment of the UNAIDs 95-95-95 goals [10]. Thus, it is essential to understand why viral load monitoring is incomplete so the gaps can be addressed.

This study showed that patient engagement in care plays an important role in the missingness of viral loads, with approximately 54.7% of patients missing their viral load because they were lost to follow up. Patients who were lost to follow up had unknown whereabouts and potentially could be out of HIV care, may have self-transferred to another clinic without documentation, or died without the clinic’s knowledge. Attrition in care engagement leads to underestimation or overestimation of the progress of HIV care and treatment programs. Participant tracing, social support, and reminder systems may improve retention and lead to more complete viral load monitoring outcomes [16]. Some studies using SMS to enhance HIV care and treatment cascade, by engaging both patients and clinicians for reminders about testing, viral load result notification, and patient tracing have proven to be effective [32], [33]. Manpower needs are critical in HIV clinics to follow up patients out of care for care engagement and viral load monitoring [34].

Our findings also revealed that patient’s missed viral load monitoring due to clinical factors. At both 12 and 24 months after ART initiation, over one third of the patients who were in care missed viral load monitoring measurements because of misinterpretation of the guidelines and failure to ensure patients sent for viral load tests in the laboratory followed through. Although the guidelines indicate viral load monitoring for adults is done at 6, 12, and 24 months after ART initiation [35], clinicians commonly interpreted the guidelines as recommending viral loads at 6 months, then 12 months from the last viral load done (e.g. 18 months) and 30 months. The study team struggled to get concurrence on our interpretation of the guidelines with some clinical and program teams during the study period. This demonstrated a gap in understanding of the guidelines translating to incomplete VL monitoring as required. Providers need to know when treatment isn’t working, and timely viral load measurement provides that knowledge. This is especially important during the first year on antiretroviral therapy (ART), as most treatment failures occur during this period [36], [37], which is why the 6 and 12-month viral load tests are recommended. It is also motivating for patients to know if their treatment is working to stay on track or to figure out what action is needed. Viral suppression allows patients living with HIV to stay health and to prevent onward HIV transmission. To mitigate these clinical challenges, capacity building through training, strong clinical guidance and support supervision by program managers are pivotal components of compliant guideline implementation [38], [39].

We observed that patients living with HIV with a WHO clinical stage 4, those in rural health facilities, younger patients <24 years and those with a higher SES had a higher risk of missing viral load tests. The advance clinical stage results are consistent with findings from a study conducted in Myanmar that examined viral load testing uptake and implementation challenges; the study demonstrated that patients with WHO clinic stage 4 had significantly higher rates of not being tested for viral load [40]. Perhaps, tracing these patients in addition to those who are loss to follow up can reduce the missed opportunity for viral load monitoring and investigating more on patients with advanced HIV. Younger patients <24 years were more likely to miss viral load measurements compared to those above 44 years. This was similar to findings from a study conducted in Gomba, Uganda looking at non uptake of VL that reported people below 25 years of age to be less likely to have a VL test [27]. Perhaps younger people are more mobile and likely to be out of care hence missing out on routine viral load monitoring [41]. Patients receiving HIV care and treatment in more rural health facilities such as Migori and Rongo counties had a higher risk of missing viral load monitoring measurements compared to those in urban facilities in Kisumu County. It has been shown that the testing rates in urban and developed health facilities are higher compared to rural areas [42]. These discrepancies are due to lack of testing equipment, poor access to health facilities, differences in the level of training of the health care workers, and weak transport and storage systems [26], [42], [43]. Low socioeconomic status has extensively been demonstrated to be associated with poor health outcomes such as increased morbidity and mortality, lack of adherence to clinic attendance and treatment [44], [45], [46], [47]. Contrary to these finding, our study showed that those who were richer/socioeconomically advantaged were at a higher risk of missing viral load monitoring measurements compared to the poorest. A possible explanation could be that they felt healthy and disregarded the viral load tests or they could possibly be among those who were disengaged in care at the time for viral load test or self-transferred to other facilities. Further investigations are needed to obtain an in depth understanding of this finding.

### Strengths and limitations

The extended outcome investigation approach is a good way to complete viral load outcomes at various clinics. Often, we do not routinely explore reasons for missed viral load measurements. This study not only examined the occurrence of missed viral loads but also explored underlying causes. The study obtained routinely collected data from the facilities thus demonstrating real world practice. The study sites’ location provided a comparison of the urban and rural facilities in Western Kenya, yet the finding may not be generalizable to other regions of Kenya. We were not able to establish other factors that could have led to missed viral load- like reagent stock outs, machine breakdown and impact of health care worker strikes on HIV care and treatment. Conducting qualitative interviews and facility assessments would have complemented our results. We had information on participants who had transferred to other health facilities but had limited outcome data. Strengthening facility linkage across clinics would ensure continuity of care and documentation of patient outcomes. Additionally, clinicians avoided viral load measurements among patients suspected to be viremic. Given that this practice is not documented, we did not obtain data on the estimates of patients who did not receive VL monitoring due to suspected viremia by the clinicians.

## Conclusions

Missed routine viral load monitoring among newly initiating PWH in Kenya remains high. Loss to follow-up is the major source of missing information and could alter overall suppression estimates substantially. Sustainable approaches to keep people living with HIV engaged and strengthening providers alignment with national guidelines can help public health systems make the most of viral load monitoring technology among patients living with HIV.

## Data Availability

Data will be available upon request made to the corresponding author

## Authors’ contributions

HAF and EG conceptualized and designed the study. HAF, SI, EN, and JK were responsible for data collection. EN performed data analysis and interpretation. HAF drafted the initial manuscript. All authors critically reviewed the manuscript, contributed to revisions, and approved the final version for submission.

## Acknowledgements

We would like to thank all the ADAPT-1 study staff for their invaluable contributions to this manuscript. Special thanks to the Research Assistants who were key in the investigation process and all the study participants.

## Data Availability Statement

The data that support the findings of this study are available on request from the corresponding author. The data are not publicly available due to privacy or ethical restrictions.

## Appendix A: Extended outcome investigation form (EOIC)

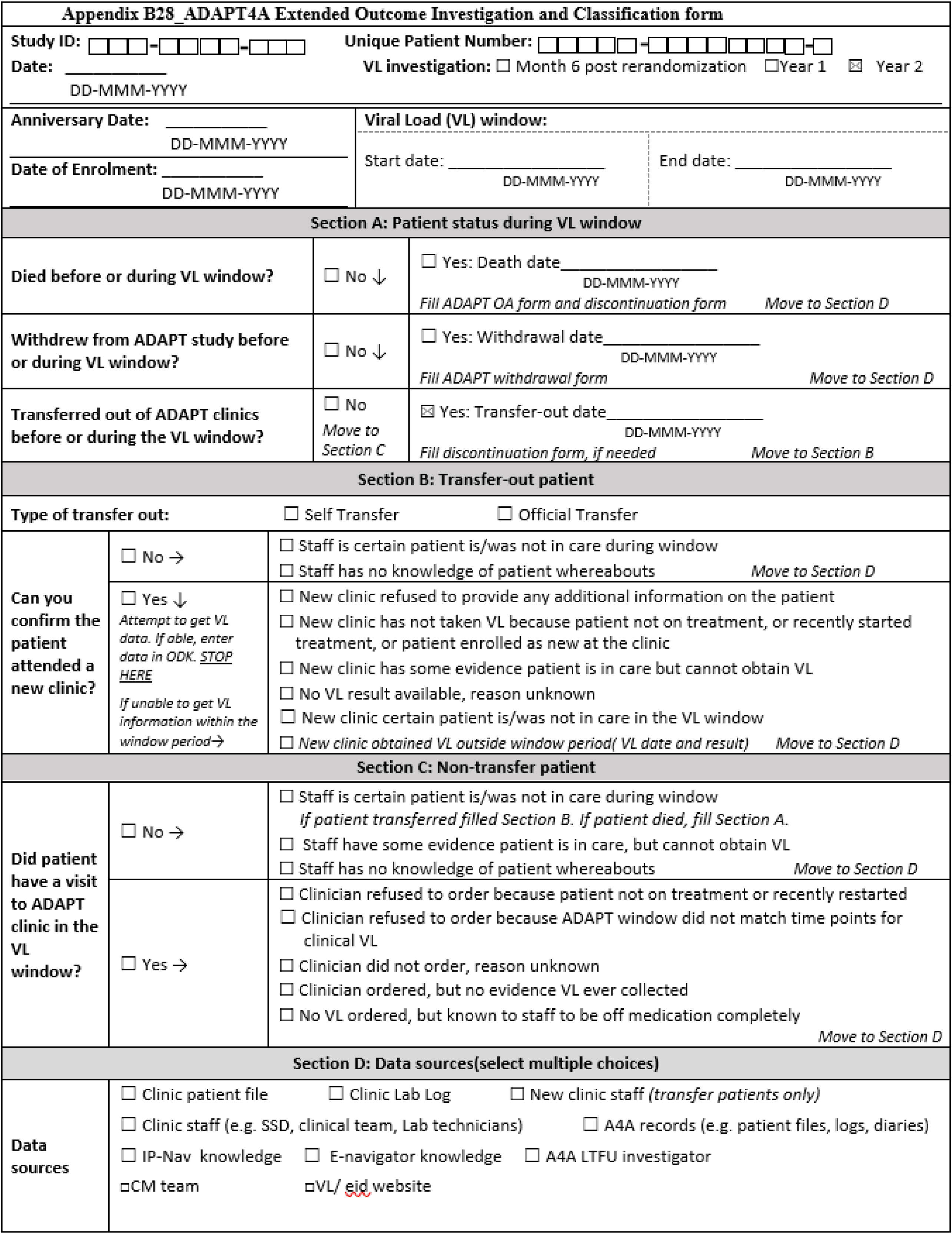

